# Evaluation of a pharmacist-led audit and feedback intervention to reduce Gentamicin prescribing errors at admission in neonatal inpatient care in Kenya: A controlled interrupted time series study

**DOI:** 10.64898/2026.01.27.26345018

**Authors:** Timothy Tuti, Jalemba Aluvaala, Mercy Mulaku, Dorothy Aywak, Muthoni Ogolla, George Mbevi, Mike English, The Clinical Information Network Group^

## Abstract

**Background:** In neonatal hospital care where the population with severe illness has a high mortality rate, around 14.9% of drug prescriptions have errors in Low- and Middle- Income Countries (LMICs) settings. However, there is scant research on interventions to improve medication safety to mitigate such errors.

**Objective:** Our objective was to explore a theory-informed pharmacist-led Audit and Feedback (A&F) intervention to improve routine prescribing practices with the aim of reducing gentamicin prescribing errors in neonatal care.

**Methods:** We used interrupted time series analysis modelling changes in prescribing errors for neonates ≤28 days admitted to newborn units (NBU) in 22 hospitals in Kenya between July 2021 to June 2024 and explored intervention effects in a feedback meeting at the end of the study. The study had three phases, pre-intervention period (July 2021 to June 2022), intervention period (July 2022 to June 2023), and post-intervention period (July 2023 to June 2024). The primary study was a standard single-group interrupted time-series study (ITS) design to evaluate the comparative effectiveness of enhanced A&F in reducing prescribing error trends after its introduction in 16 hospitals. Secondary analysis included comparison to prescribing error outcomes in an additional six hospitals in a contemporaneous control group that received basic A&F reports without pharmacist involvement in the NBU prescribing practices.

**Results:** Between July 2021 and June 2024, the 16 hospitals in the primary outcome analysis and the 6 additional hospitals for the secondary outcome analysis had 36,668 and 8,943 neonates with Gentamicin prescriptions at admission retrospectively. From the incidence rate ratios (IRR) of incorrect prescribing at admission, there was no step change (IRR 1.115, 95% CI: 0.920 to 1.352, p-value=0.265) or trend change (IRR 1.014, 95% CI: 0.986 to 1.042, p-value=0.344) due to the enhanced pharmacist-led A&F intervention in the 16 hospitals in the primary study. From the secondary study, change in the trend post-intervention in the 16 primary study hospitals in the primary study relative to the 6 hospitals acting as a contemporaneous control group was positive (IRR 0.933, 95% CI: 0.878 to 0.985, p-value=0.014), despite no step change due to the enhanced A&F intervention.

**Conclusion:** We found no statistically significant effect of the team-based pharmacist-led A&F intervention on reducing gentamicin medication errors in neonatal care. Prescribing errors during intervention and post-intervention periods were increasing across all hospitals in both arms of the study during and post-intervention periods. However, relative to control hospitals sites receiving routine feedback but without pharmacist involvement or pharmacist-led CMEs, the primary study sites had a positive trend in reducing Gentamicin prescription error rates at admission during and post-introduction of the pharmacist-led A&F intervention.

**Trial registration:** *PACTR*, *PACTR202203869312307.* Registered 17^th^ March 2022, https://pactr.samrc.ac.za/Search.aspx?TrialID=PACTR202203869312307

**Why was this study done?:**

- In newborn hospital care where the population with severe illness has a high mortality rate, around 14.9% of drug prescriptions have errors in settings such as sub-Saharan Africa (SSA).
- However, there is scant research in SSA on actionable audit and feedback interventions over time to reduce the rates of inappropriate and potentially harmful prescribing of antibiotics.
- Therefore, we evaluated whether such an intervention is associated with sustained changes when it provides continuous feedback championed by pharmacists.

**What did the researchers do and find?:**

- We evaluated the impact of a pharmacist-led audit-and feedback intervention for in-hospital newborn care across Kenya.
- We found that the intervention was not associated with sustained reduction in the level or trend in incorrect antibiotic prescribing across practices, until the study was completed (after 12 months).
- Despite the overall increase in prescribing errors during the study period and the 12 months after the study period, a marked difference in inaccurate prescribing trend was also seen between hospital groups where the hospital pharmacist agreed to be involved with the audit and feedback intervention.

**What do these findings mean?:**

- The extent to which actionable audit and feedback interventions reflect the complexity of routine hospital care in SSA determine whether long-term improvements in prescribing practices can be delivered on an ongoing basis.
- More research is needed to understand why and how to obtain sustained reductions in antibiotic prescribing errors during hospital stay in SSA.

## Introduction

Improving medication safety is a global priority as medication errors arising from prescribing, dispensing, transcribing, administering, and monitoring medicines can cause severe harm and increase healthcare costs [1–3]. Most evidence on medication safety in routine healthcare settings is from high-income countries (HICs) [3]. From the limited findings available, drug prescribing errors might be substantively higher in low- and middle-income countries (LMICs) [4–6], especially as electronic prescribing with in-built error checking remains very limited in many public hospitals in LMICs as part of limited adoption of Electronic Health Records (EHRs) [7, 8].

The challenge of poor medication prescribing practices in hospital care is especially pertinent in the neonatal period (i.e. first 28 days of life) [9, 10] where the population with severe illness has high mortality [11], and dose calculations are often complex. Around 15% of drug prescriptions have errors in neonatal care settings [9], but there is scant research on interventions to improve medication safety in LMICs where neonatal sepsis is common [12] and where concerns over antibiotic resistance and antimicrobial stewardship are growing [13].

In this study, we explored a pharmacist-led, audit and feedback (A&F) intervention guided by the Clinical Performance Feedback Intervention Theory (CP-FIT) to <u>R</u>educe <u>Ge</u>ntamicin dosing errors in <u>N</u>ewborn <u>T</u>reatment (i.e. ReGENT study) in Kenyan hospitals. We build on previously reported studies [14–16] and explain the intervention in detail in this study’s published protocol [17]. Here we report the evaluation of this pharmacist-led A&F intervention approach and its effects on the prevalence of gentamicin prescribing errors in neonatal inpatient hospital care over a three years period.

This study’s objective was to assess whether an enhanced pharmacist-led audit and feedback intervention reduces neonatal gentamicin prescribing errors more effectively than standard hospital feedback over time.

## Design and methodology

### Ethics approvals

The analyses described in this study were approved by the KEMRI’s Scientific and Ethical Review Committee (SERU #4378 and SERU #3459) with detailed information provided in the protocol [17]. Our description follows the TREND statement [18] for improving the reporting quality of non-randomised evaluations of interventions (Appendix) and the Template for Intervention Description and Replication (TIDieR) Checklist [19] (Appendix).

### Study setting

The study was conducted in partnership with 22 hospitals in Kenya. These hospitals were purposefully selected to join a Kenyan Clinical Information Network (CIN, see below) [8, 20, 21], be of at least moderate size and representative of different malaria transmission zones. These hospitals admit small and sick newborns to a New-Born Unit (NBU) with a specific clinical and nursing team. The average age of neonates on admission is 0 or 1 day old and most admitted neonates are inborn [20]. These hospitals joined the CIN, a learning health system in Kenya between 2014 and 2020 [8, 20, 21]. The hospitals already receive 3 monthly clinical audit and feedback reports on the quality of care they provide for common conditions, which include a summary of prescription error rates for gentamicin and penicillin [22]. The quarterly report is shared to all CIN sites via email and as a printed copy to the paediatrician and the hospital manager, providing a common A&F intervention already in use. Neonatal team leaders (neonatologists, paediatricians, and nurses) also met face-to-face once or twice annually to discuss these standard A&F reports and how to improve multiple facets of clinical care [23, 24]. In these hospitals, in Kenya more widely and in many public sector settings in LMICs, key issues that likely influence prescribing practices and affect dose errors, paying special attention to gentamicin the first-line antibiotic for severe neonatal infection, are listed in table 1.

**Table 1:**
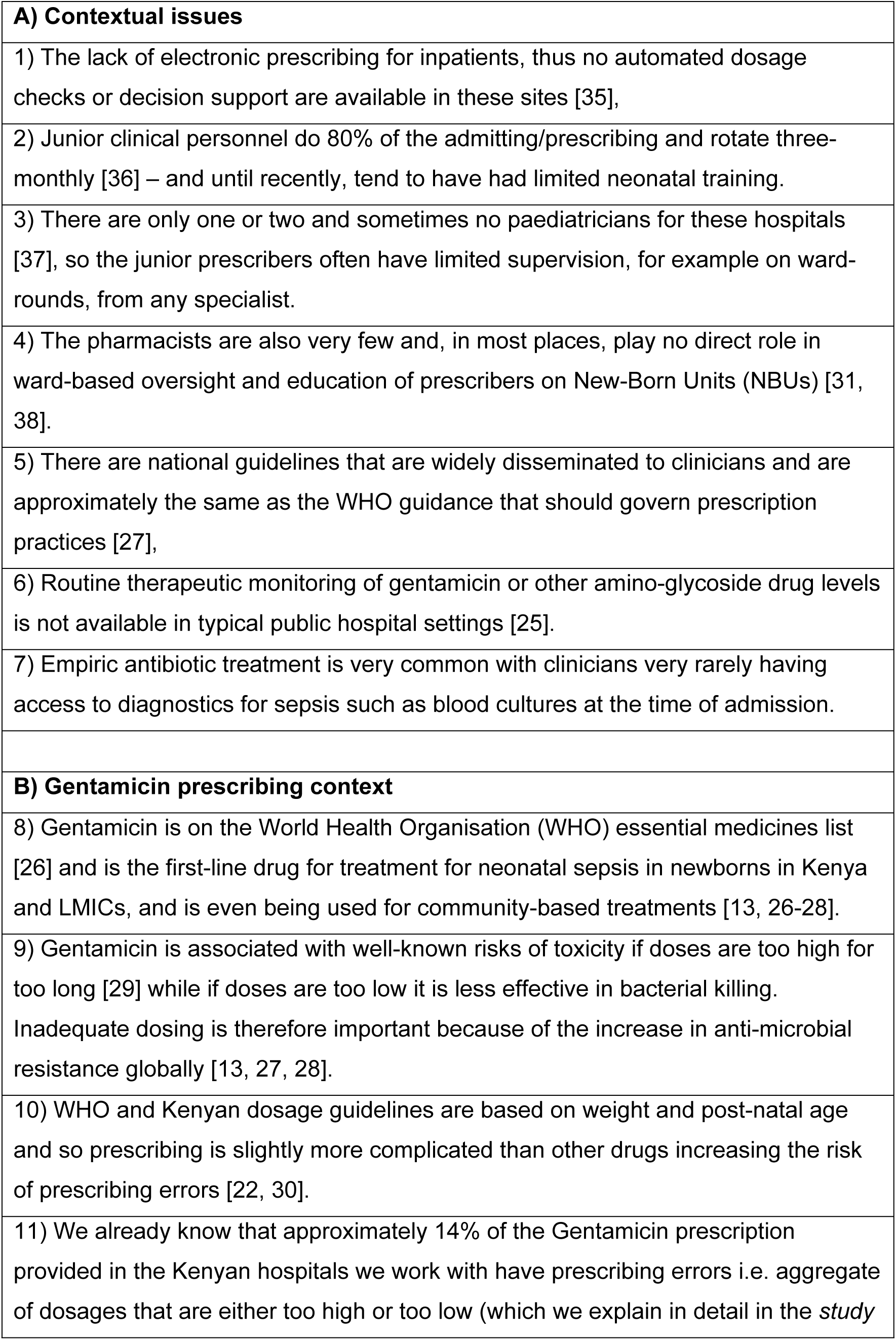
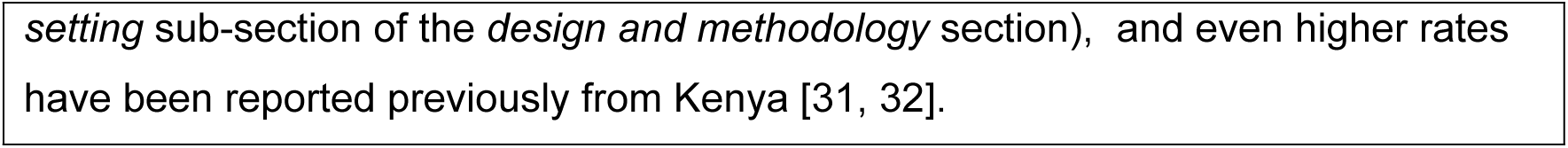
Contextual and Gentamicin prescribing factors in typical neonatal units in public health facilities in LMICs.

### Study design

The study design is described in detail in the published protocol [17]. In summary, the primary study was a standard single-group interrupted time-series study (ITS) design for the primary analysis to evaluate the comparative effectiveness of the enhanced A&F in reducing prescribing error trend after its introduction (which we explain in detail in the intervention section). No randomisation was planned, the aim was to include all hospitals in the intervention but pharmacists from 6 CIN facilities refused to participate [17]. This provided an opportunity for an intervention versus control comparison as a secondary analysis using a parallel group ITS design to evaluate prescribing error trends in the two hospital groups (Figure 1) while recognising the potential selection bias in the facilities that rejected the pharmacist-led enhanced A&F intervention approach.

**Figure 1:**
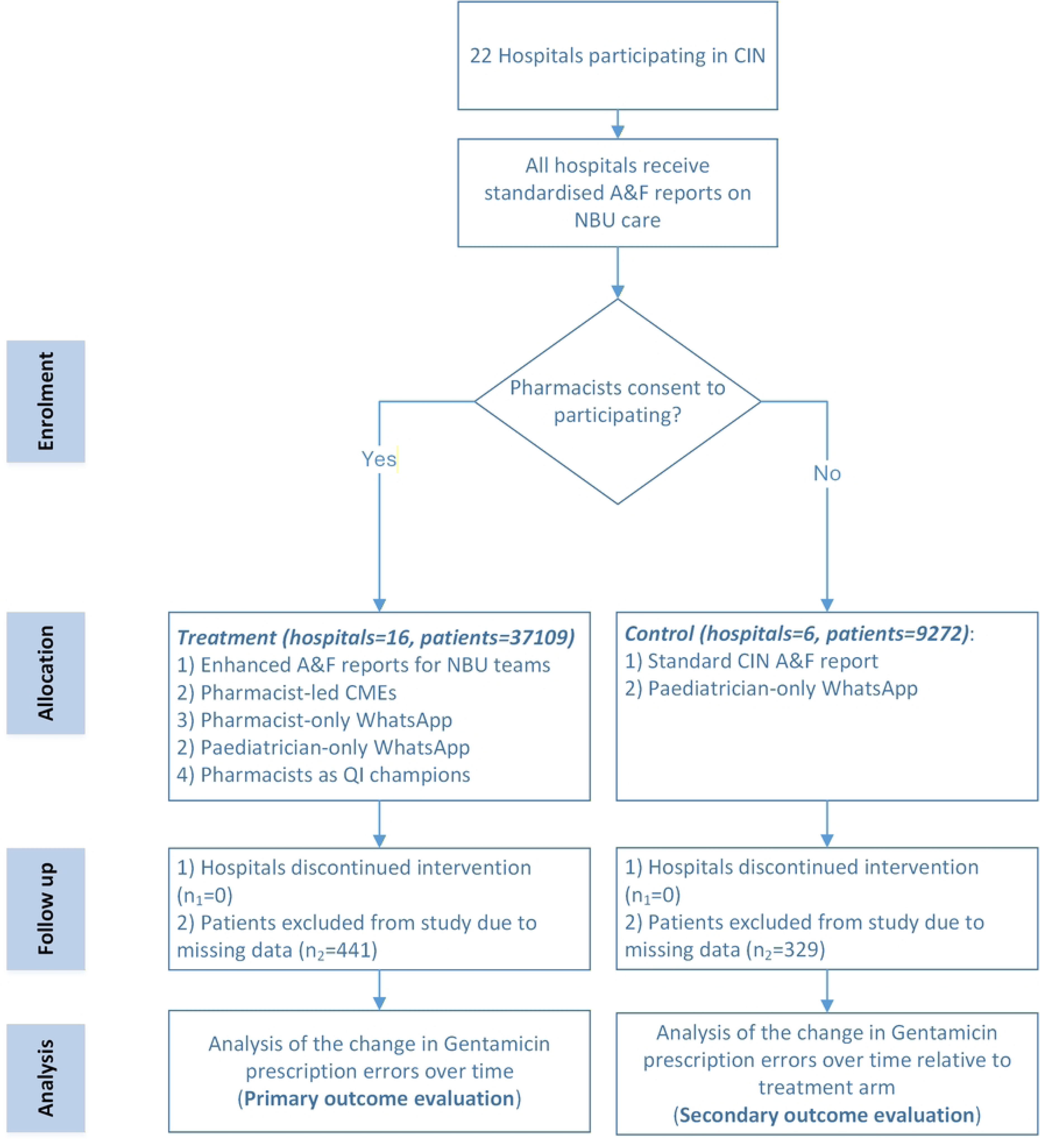
Flow chart of the intervention roll-out. The ITS starts prior to hospitals being assigned to control or experiment group

### Intervention: Audit and Feedback (A&F)

A&F is posited to be more effective when it has multiple components (e.g., education), involves key agents (facilitators and champions) such as pharmacists, and addresses individual and team goals [14, 33]. The roles of key agents (facilitators and champions), for example pharmacists, for prescribing practices improvement have only rarely been empirically tested [34, 35]. Feedback directly to clinicians on their performance is posited to be most effective [16] but healthcare workers (HCWs) are few in number in many LMICs with a finite supply of time and resources to individually engage with feedback [16].

In many LMIC hospital settings, the type of clinical data and digital platforms available in routine clinical settings constrain the nature of A&F intervention. A&F studies need to consider what advantages might be gained by leveraging local clinical champions, and the value and practicality of feedback at team or individual levels [16]. The evaluation of the effect of A&F interventions also needs to consider the decay in effect over time by aiming to go beyond a single feedback cycle, short-term intervention period or simple before-after designs [36].

In response to calls for more ‘head to head’ studies of different forms of A&F to help identify ‘what works’ [16], our A&F design choices took account of both the specific characteristics of these contexts and existing knowledge and theory [37, 38] in the following ways:

1. The feedback visualisations were limited to three and delivered as a one-pager PDF with infographics to reflect HCWs finite capacity to handle yet another feedback report. These were designed to communicate explicit goals within the control of the recipients and whether clinical team performance had room for improvement, and allow benchmarking with peer hospitals, while improving the specificity and trend reporting of performance [16].
2. We provided an Android smartphone-based feedback performance dashboard to ensure active A&F delivery and ease automated performance analysis and instantaneous presentation of feedback to recipients [16].
3. Because HCWs have strong beliefs regarding how care should be provided which in turn influences their interactions with feedback, the provision of a WhatsApp avenue was provided to help facilitate and evaluate if the perceptions, acceptance, and intentions teased from their interaction with the A&F intervention align with the observed clinical behaviours [16].
4. We targeted the use of pharmacists as champions in A&F strategy because where HCWs with source knowledge and skill advocated for the feedback intervention, they often influenced others and provided additional resource to enable more meaningful engagement with feedback [16].
5. We provided pharmacist-led Continuous Medical Education (CME) seminars linked to NBU antimicrobial stewardship and prescribing practices (Supplementary Table 1) to strengthen both the knowledge and skills in quality improvement and in the AMS clinical topics [16].

The theoretical underpinnings of the enhanced A&F report visualisations are provided in detail in the protocol [16, 17]. The enhanced A&F feedback reports in this study provided feedback through: (1) score cards reporting deviation from explicit targets, (2) peer comparison of Gentamicin prescribing errors by newborn patient sub-groups, and (3) hospital-specific performance trends over a six-month period summarised by newborn age-groups [17]. The differences in the A&F components between the study arms are illustrated in Figure 1. Details of the feedback components, together with the enhanced A&F PDF infographics are provided in Appendix 1’s Supplementary Table 1-2 and the published protocol’s Supplementary Figures 1-3 [17].

### Outcomes

The primary outcome of this study was the proportion of patients in newborn units receiving an inaccurate Gentamicin prescription. The calculation of the correct prescription according to the Kenyan guidelines is age and weight dependent as illustrated in figure 2 where Gentamicin prescription is reported in milligram units. In summary, for neonates <7 days old, the correct gentamicin dosage is 4 mg/kg to 6 mg/kg, or 2.4 mg/kg to 3.6 mg/kg if birth weight is less than 2000 grams. For neonates who are 7 days old and older, the correct gentamicin dosage is between 6 mg/kg and 9 mg/kg (Figure 2).

**Figure 2:**
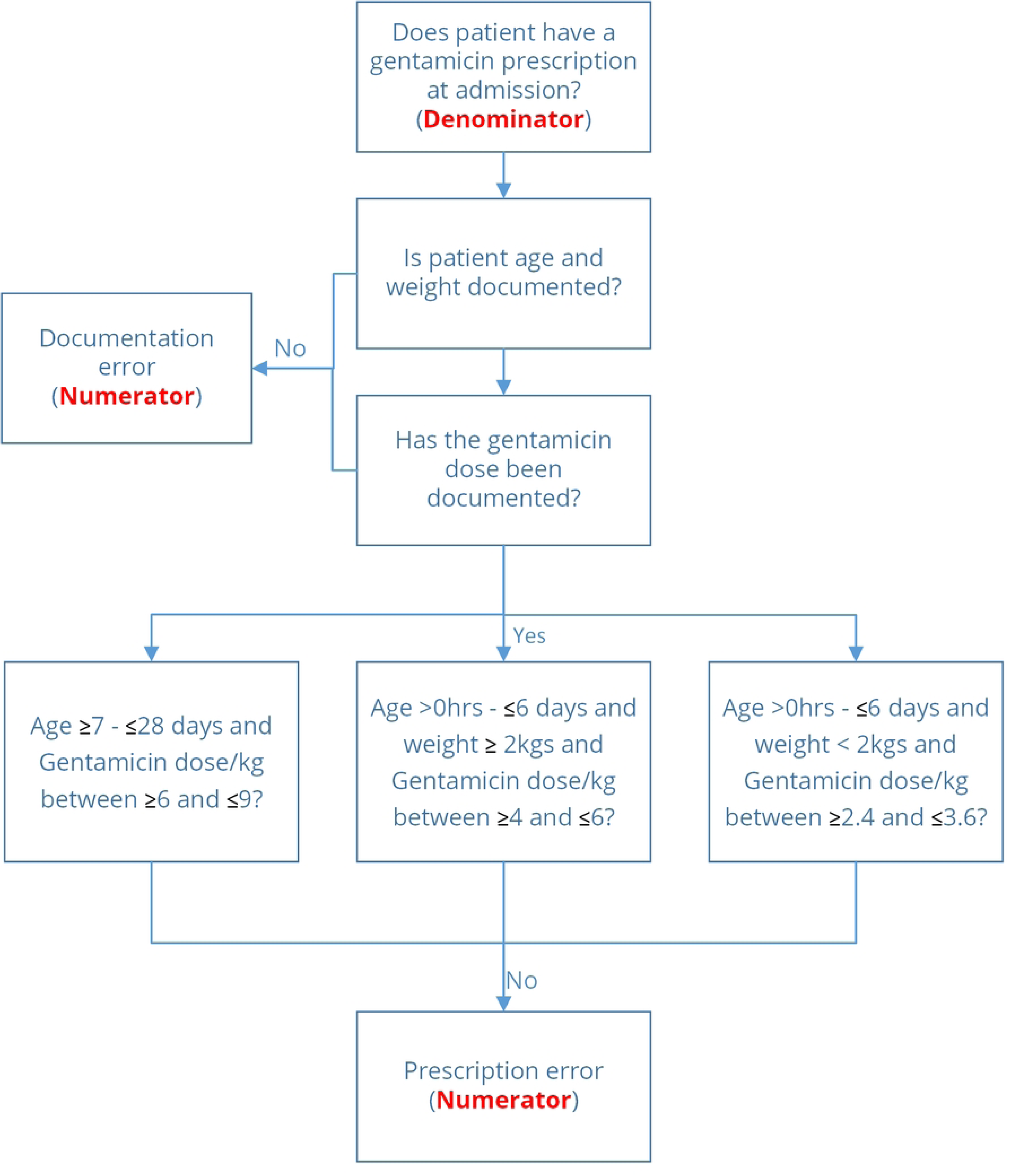
Study primary outcome from patients admitted to the NBUs. Dosage calculations per kilogram allow for ±20% deviation, outside which they are considered errors.

From the CP-FIT model, this outcome represents the standard of clinical performance against which clinical behaviour would be measured explicitly (*Goal setting*) [16]. This study’s target process outcome was at least a 35% reduction in prescription errors from individual hospital baseline error rates with this target based on published evidence of 35-50% reductions in prescription errors in neonatal care from before-after studies, which translates to an absolute change from 14% to 9.1% in prescribing error reduction [35].

### Data collection procedures and management

Methods of collection and cleaning of data in the CIN are reported in detail elsewhere [22, 39]. In summary, clinical data for neonatal admissions to the hospitals within the CIN were captured through structured Neonatal Admission Record (NAR) forms coupled with standard treatment sheets that are approved by the Ministry of Health. The NAR prompts the clinician with a checklist of fields including patient biodata, clinical assessment, admission and discharge diagnoses, and record of outcome (survival or death). The CIN supports one data clerk in each hospital to extract data from paper medical records, nursing charts, treatment charts, and available laboratory reports each day after a newborn’s discharge into the primary data collection tool developed in Research Electronic Data Capture (REDCap). Automated error checking happens at the point of entry by daily review, every week centrally and both are complemented by regular external data quality assurance reviews [39]. A minimal dataset – which is unsuitable for our planned analyses - is collected for (1) admissions during major holiday breaks, (2) admissions when the data clerk was on leave, and (3) on a random selection of records in hospitals where the workload is very high. This process is explained in detail elsewhere [39, 40]. All data in de-identified form derived from medical records was stored in secure KEMRI-Wellcome Trust Research Programme servers with specified researchers provided password protected access.

For this study deidentified data from all patients admitted to the selected hospitals’ NBUs who are under the age of 28 days (i.e., neonates) who had a Gentamicin drug prescription on admission were eligible for inclusion in the analysis regardless of gestational age as in Kenya prescriptions are based on weight and postnatal age rather than gestation at birth. Only the patient population with full data on age, weight, and the dose of the prescription were included in the outcome measurement since these data are needed to measure errors. Inadequate Gentamicin prescription documentation occurred in <1% of all prescriptions documented within CIN in the 24 months before intervention introduction. During the intervention period and the analysis stage, the participating hospitals and clerks capturing prescribing data were blinded to the study arm assignment, but the research staff were not.

### Protocol deviations

In the 16 facilities that received the intervention, contrary to the expectations in the published protocol, none of the pharmacists were willing to join the NBU clinical team WhatsApp group where one existed or form a NBU WhatsApp group where none existed. All healthcare workers save 6 did not use the mobile application to access the performance dashboards, preferring the monthly PDF reports instead. These deviations rendered no substantive difference in the enhanced A&F intervention packages between the intended study arms as envisioned in the published protocol (Table 1, Figure 1)[17]; We still provide analysis results based on the intention-to-treat principles.

Post-hoc analysis of the 12 months after the intervention had ended was decided upon at the end of the intervention roll-out to evaluate whether any intervention effect would be sustained after the study by comparing the 16 sites that had received the pharmacist led A&F intervention versus the 6 sites that only receiving the standard A&F reports from the CIN.

### Data analysis and statistical methods

This study involved all healthcare workers prescribing gentamicin in the NBUs of the participating hospitals. On average, there is one paediatrician, one medical officer and several medical officer and clinical officer interns who might prescribe. They are supported by three daytime nurses, and two night-nurses in a typical NBU unit. As medical officer and clinical officer interns rotate typically every 3 months the total number of prescribers working in the NBU units over the period was in the hundreds [41] making it difficult to attribute a patient’s Gentamicin prescribing to individual clinicians.

### Interrupted Time Series analysis

We applied a segmented linear mixed effects model with an autoregressive covariance structure on the proposed Interrupted Time Series (ITS) design, that accounts for the pre-intervention trends in the study outcomes [42]. Informed by previous findings [35], our hypothesised impact model assumed month-to-month (slope) changes following the implementation of the intervention, but no immediate (level) change. We adopted a negative binomial modelling approach which we explain in detail in the protocol [17], and report study outcomes as incidence rate ratios (IRR), where values with 95% confidence intervals(CI) >1 indicate increase in prescribing error rates while the reverse (i.e. 95% CI < 1) indicate decrease in prescribing error rates at admission; IRR values with 95% CI containing 1 indicate no change in the prescribing error rates at admission. The primary analysis for this study was the comparison of pre-intervention to post-intervention trends of the study outcomes in the facilities that consented to participate in the enhanced A&F intervention approach. We also conducted secondary analysis comparing the pre- and post-intervention trends in the study outcome between the facilities that were on the enhanced A&F approach compared to those that were on the basic A&F approach.

### Interrupted Time Series (ITS) Sample size

Detailed explanations on our approach to sample size calculations are provided in our published protocol [17]. The primary analysis followed a single group ITS study design in keeping with a priori study design and entailed the 16/22 facilities that consented to pharmacist-led enhanced A&F intervention approach. All analysis was done in R (v 4.4.3) statistical computing software [43].

### Primary analysis

The baseline level of erroneous Gentamicin prescribing at admission was 12.07% (95% CI: 11.56% - 12.58%) across the intervention sites (n=16) from the pre-intervention period. From published reviews, A&F interventions have a median absolute effect size of 4.3% in improving practice behaviour; We estimated that with 950 patients per month across the 16 facilities, our study would have 80% power to detect this 4.3% reduction of the month-to-month prescription error trend over 12 months (i.e. slope change) with a statistical significance of 0.05. The monthly average of admissions to NBUs with a gentamicin prescription at admission during the pre-intervention period in all the 16 facilities that consented to the enhanced pharmacist-led A&F intervention approach was 1325 patients (with a standard deviation of 70 across the 16 hospitals).

### Secondary analysis

The 6/22 CIN sites that chose to not participate in the study were treated as a control group within the ITS design to examine whether improvements in performance (correct prescribing) were more pronounced than improvements that may be linked to underlying secular trends [44]. When considering both the intervention arm and control arm (sites = 22), the baseline monthly level of erroneous Gentamicin prescribing at admission was 12.3% (95% CI: 11.84% - 12.76%) across the 22 facilities from the pre-intervention period. Based on previously published studies [35], we estimated that with 835 patients per month across the 22 facilities, our study would have 85% power to detect an effect size of 15% reduction of the month-to-month prescription error trend over 12 months (i.e. slope change due to arm) with a statistical significance of 0.05. The monthly average admissions to NBUs with a gentamicin prescription at admission in all the 22 facilities during the pre-intervention period was 1617 patients (with a standard deviation of 76 across the 22 hospitals).

Sample size analysis at the individual hospital level revealed that all the hospitals did not have sufficient patient numbers per month with gentamicin prescription to facilitate separate within hospital time-series analysis.

### Process evaluation to assess fidelity

We used messages shared over the intervention period in the study’s pharmacists WhatsApp group (which was specifically designed as a group for intended intervention champions) to explore group members’ engagement with the enhanced A&F intervention during the study period. The messages shared were summarised according to their source, target, timing, and content. While the content of the shared messages did not lend itself to the exploration of the reception, comprehension, and acceptance of the feedback by the pharmacists (*Interaction*, *Perception*, and *Acceptance* respectively) and planned behavioural responses that may be attributed to the feedback (*Intention* and *Behaviour*), we qualitatively summarised the messages for content on barriers to behaviour change linked to the A&F intervention [16]. We also convened a feedback meeting with the HCWs in the intervention sites to explore challenges and successes from the intervention implementation in CIN hospitals and to learn and gather feedback on how to better design and implement future A&F interventions.

## Results

### Participant flow and recruitment

The study’s first phase which involved baseline data collection 12 months prior to intervention introduction ran from 01^st^ July 2021 to 30^th^ June 2022 (pre-intervention period). The intervention phase ran for 12 months afterwards from 01^st^ July 2022 to 30^th^ June 2023. The post-intervention period ran for an additional 12 months from 01^st^ July 2023 to 30^th^ June 2024. Of the 46381 patients from the 22 CIN hospitals prescribed Gentamicin at admission and eligible for inclusion in this study, 37109/46381 (80.39%) of patients from 16/22 hospitals were in the arm that received the enhanced A&F intervention (Figure 3), while 9272/46381 (19.99%) were from 6/22 hospitals in the control arm. While no hospitals were lost to follow-up, 441/37109 (1.19%) and 329/9272 (3.55%) patients from the experiment and control arm were dropped due to missing data on key prescribing variables of age and weight (Figure 1, Figure 3). Subsequently, 36668/37109 (98.81%) and 8943/9272 (96.45%) of the patients were in the experiment and control arm respectively were included in the subsequent analysis.

**Figure 3:**
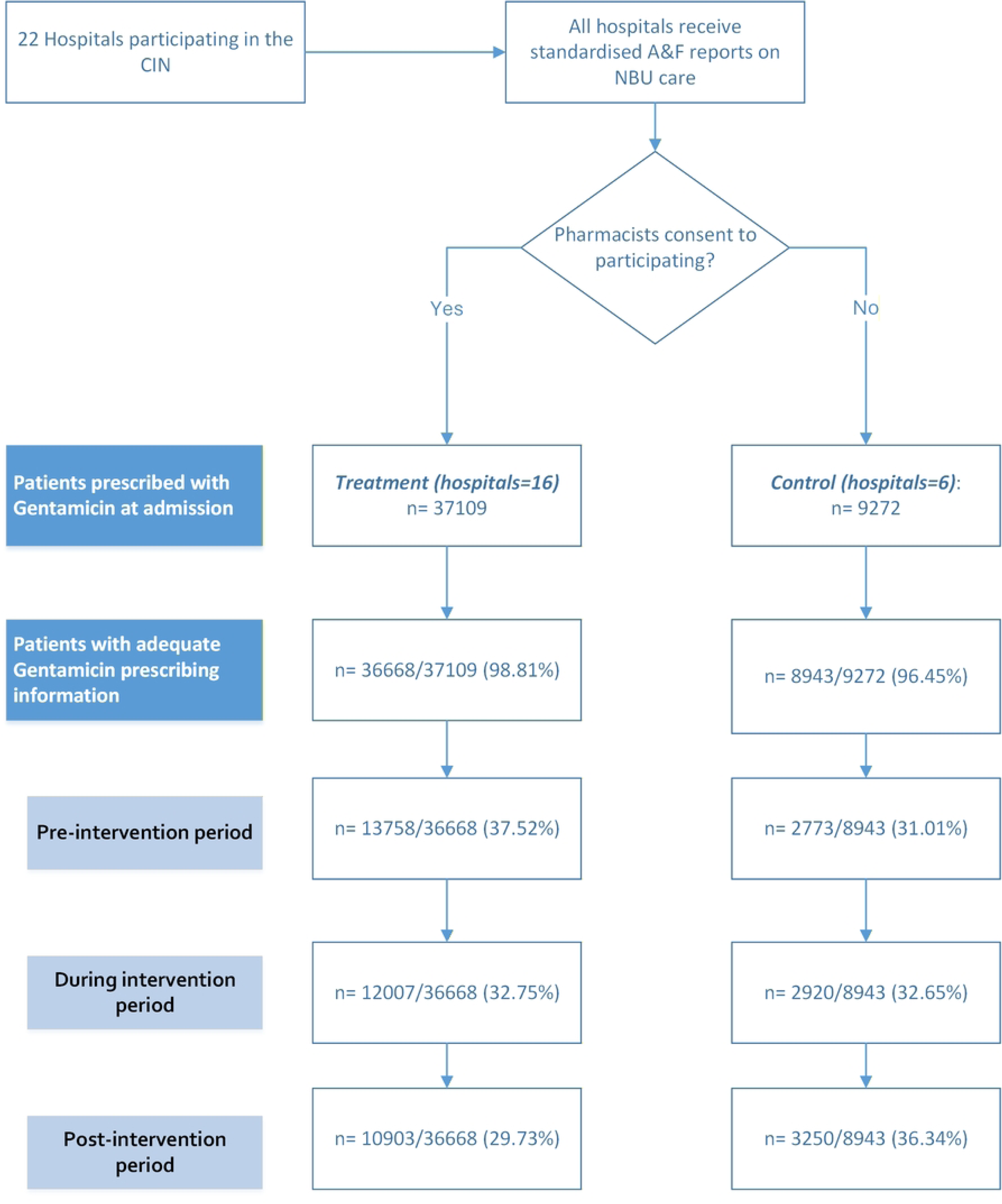
Flow-chart of patients included in the ReGENT study analyses

### Baseline data and numbers analysed

Table 2 illustrates the baseline demographic and clinical characteristics for patients in each arm of the study. Patients in both arms had similar gestations, age, birth weight, length of hospital stay and mortality rate despite the large difference in the number of patients in the study arms. From summary descriptive analysis of before and after the intervention, the gentamicin prescribing accuracy after the enhanced A&F intervention was introduced appears no different in the experiment arm compared to the period before the intervention (Figure 4), the same pattern appears true for the control arm. Figure 4 also provides the absolute error rates by patient sub-group which are important results that are rarely reported from SSA.

**Figure 4:**
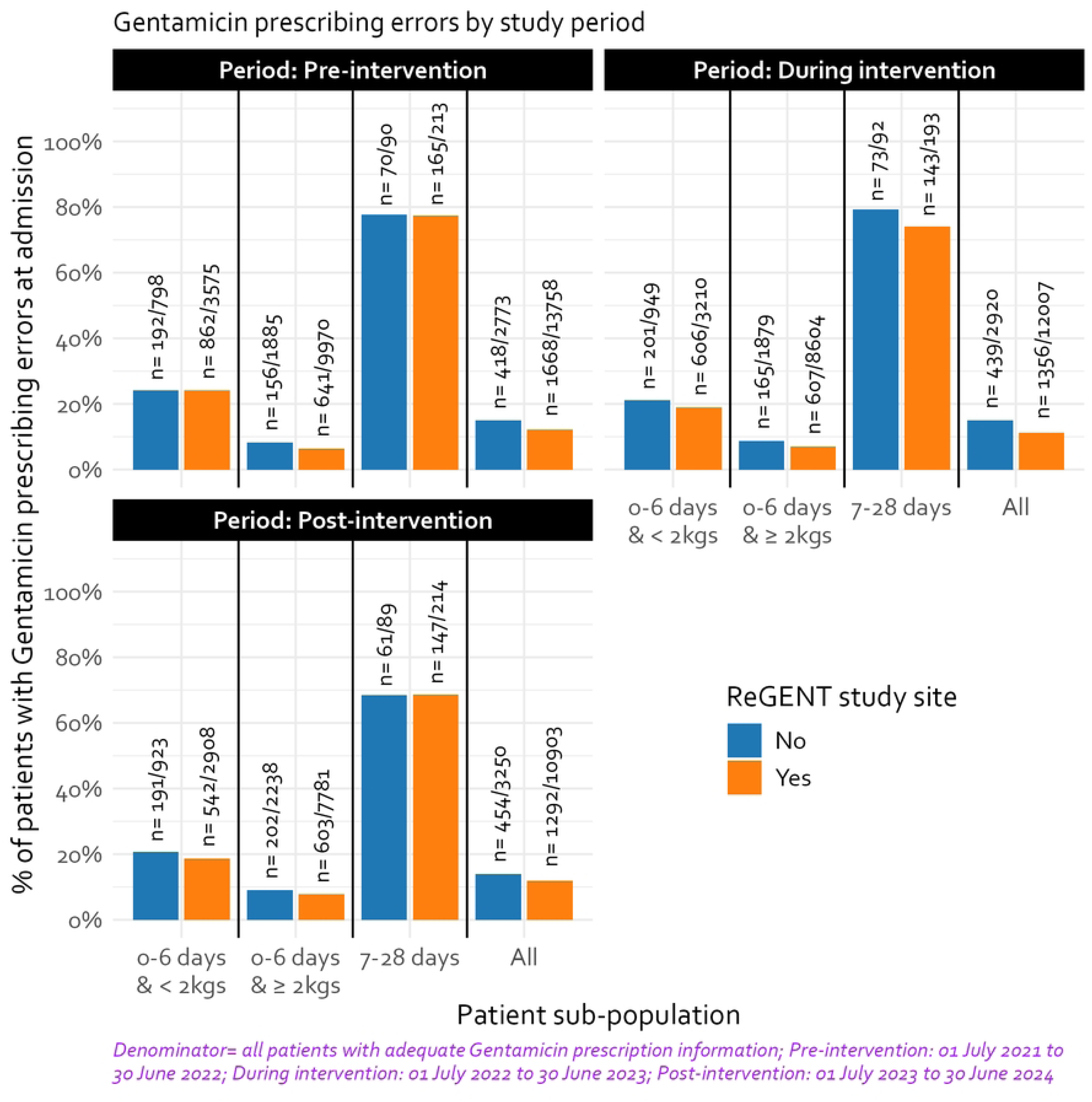
Breakdown of the Gentamicin prescribing errors at admission by study period and ReGENT study participation

**Table 2:** Descriptive Summary Statistics (n = 45611)

| Table 2: Descriptive Summary Statistics (n = 45611) |  |  |  |  |
| --- | --- | --- | --- | --- |
| Indicator | Control Arm, n= 8943 |  | Experiment Arm, n= 36668 |  |
|  | Summary Statistic<sup>1</sup> | Missing Rate | Summary Statistic<sup>1</sup> | Missing Rate |
| Neonate Age (Days) | 1 (IQR: 1-1) | 0 (0%) | 1 (IQR: 1-1) | 0 (0%) |
| Birth Weight | 2.65 (IQR: 1.81-3.2) | 0 (0%) | 2.7 (IQR: 1.9-3.2) | 0 (0%) |
| SENSS Score<sup>2</sup> | 7.6 (IQR: 3.9-16.8) | 0 (0%) | 7.1 (IQR: 3-14.7) | 0 (0%) |
| Gestational Age<sup>3</sup> | 37 (IQR: 33-39) | 826 (9.24%) | 38 (IQR: 34-39) | 3179 (8.67%) |
| Male | 5032 (56.27%) | 27 (0.3%) | 20774 (56.65%) | 87 (0.24%) |
| Length of stay (Days) | 6 (IQR: 3-12) | 18 (0.2%) | 5 (IQR: 3-10) | 100 (0.27%) |
| Mortality Rate | 1468 (16.42%) | 14 (0.16%) | 5411 (14.76%) | 18 (0.05%) |
| **Note:**<br><sup>1</sup> Continuous variable summaries given as median (Interquartile range). Discrete variable summaries given as proportions<br><sup>2</sup> SENSS (Score of Essential Newborn Symptoms and Signs) score provided as a mortality risk probability (derived from sex, birthweight, presence or absence of difficulty feeding, convulsions, indrawing, central cyanosis and floppy/inability to suck).<br><sup>3</sup> Provided in weeks |  |  |  |  |

The trends in Gentamicin prescribing errors in the experiment arm was less variable compared to the control arm for both overdose and underdose errors (Figure 5). The prescribing error trends were also quite variable in both study arms for the different neonatal inpatient sub-populations (Supplementary Figure 1-2). There is a strong indication of a large between-hospital variability in the prescribing error trends in both study arms (Supplementary Figure 3-4).

**Figure 5:**
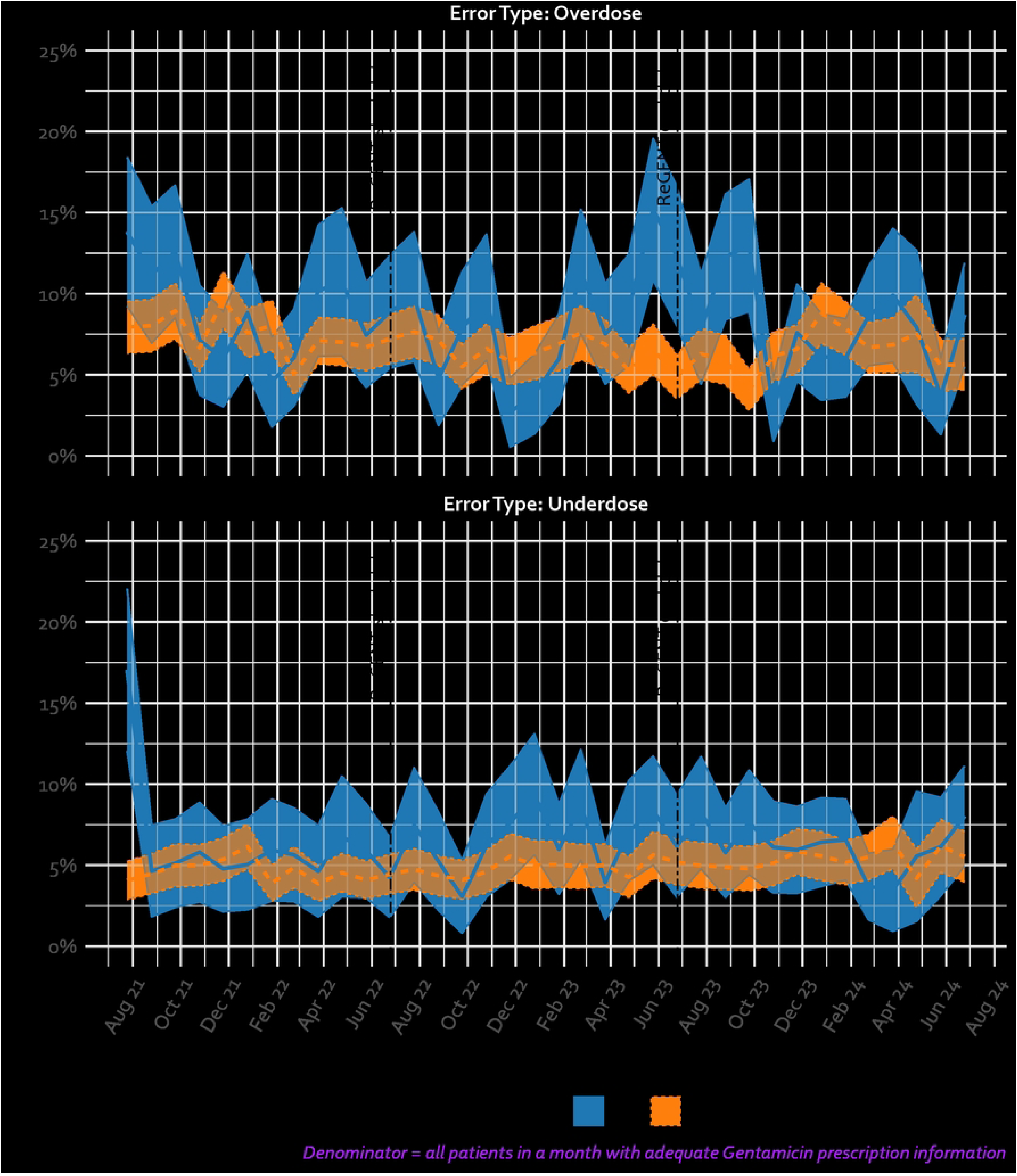
Overall trends in Gentamicin prescribing error at admission

### Outcomes and estimation

#### Intervention effect in the enhanced A&F group

Overall, from the intention-to-treat analysis results (Supplementary Table 3), there was no enhanced A&F intervention effect to reduce the incidence of inaccurate Gentamicin prescription in neonates at admission (Table 3, Figure 6), either as a step or trend change. This may be a consequence of very limited fidelity to the intervention as specified in the protocol [17].

**Figure 6:**
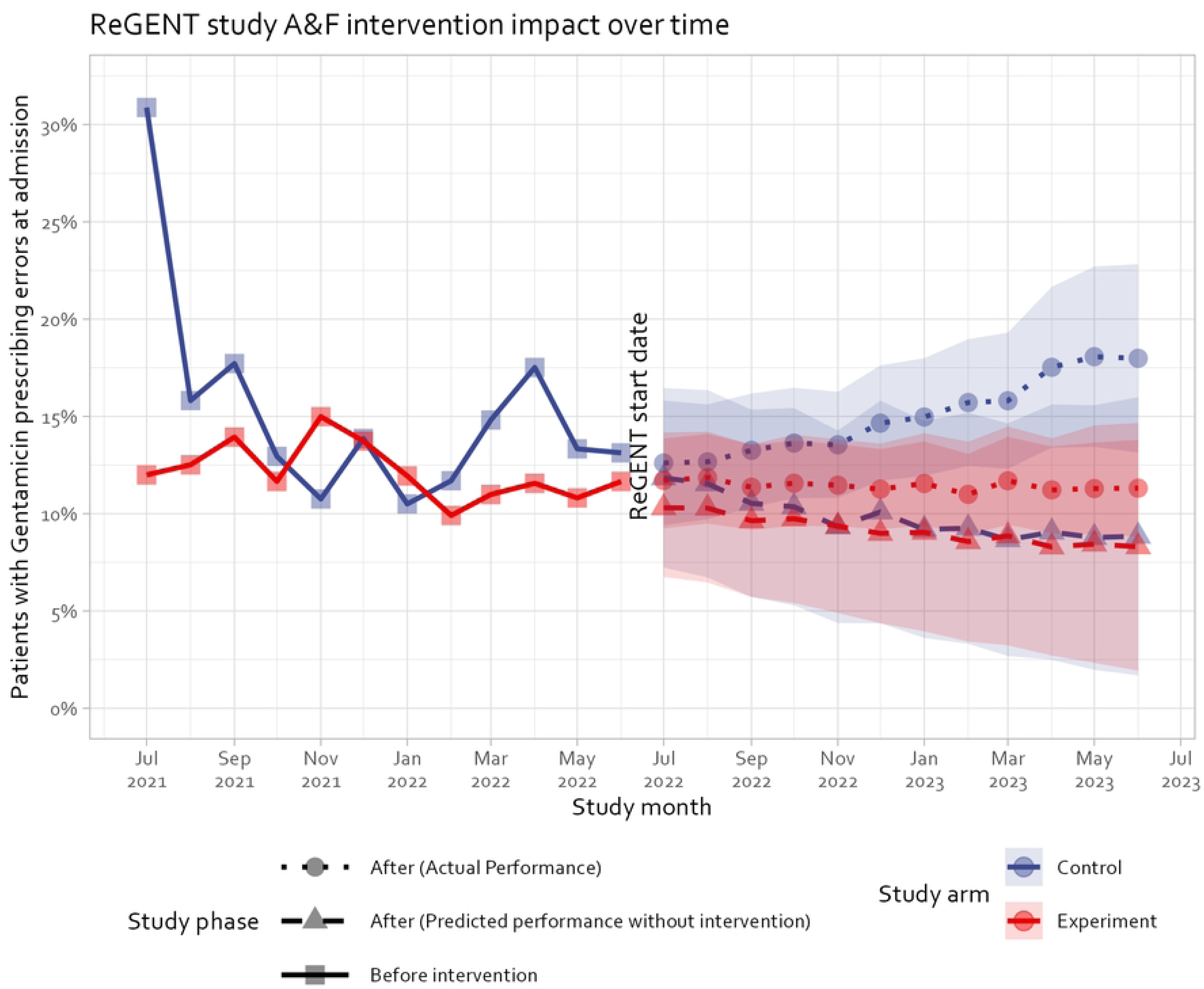
The modelled impact of ReGENT study on all Gentamicin prescribing errors at admission

**Table 3:** ReGENT study intervention effect on reducing all Gentamicin prescribing errors at admission (Hospitals = 16)

|  |  |  |  |  |  |  |
| --- | --- | --- | --- | --- | --- | --- |
|  | Pre-intervention phase compared to intervention phase |  |  | Intervention phase compared to post-intervention phase |  |  |
|  | IRR | 95% CI | p-value | IRR | 95% CI | p-value |
| (intercept)<sup>1</sup> | 0.158 | 0.119 to 0.210 | <0.001 | 0.145 | 0.099 to 0.211 | <0.001 |
| time<sup>2</sup> | 0.980 | 0.962 to 0.999 | 0.043 | 0.986 | 0.945 to 1.030 | 0.531 |
| intervention<sup>3</sup> | 1.115 | 0.920 to 1.352 | 0.265 | 0.890 | 0.695 to 1.138 | 0.352 |
| intervention × I(time - 12)<sup>4</sup> | 1.014 | 0.986 to 1.042 | 0.344 | 1.044 | 0.964 to 1.131 | 0.289 |
| **Note:**<br><sup>1</sup>Baseline performance at the start of the pre-intervention period<br><sup>2</sup>Pre-intervention trend<br><sup>3</sup>Step change in performance due to the intervention<br><sup>4</sup>Change in the trend post-intervention |  |  |  |  |  |  |

In the 12 months post-intervention for the 16 hospitals receiving the enhanced A&F intervention, the trend in gentamicin prescribing errors at admission worsened, increasing by 4.4% (p-value=0.289) month-to-month, although it was not statistically significant (Table 3).

#### Secondary analysis: Intervention effect in the enhanced A&F group in comparison to the control arm

Relative to the control arm, the experiment arm had a 7.3 % (p-value=0.009) reduction in prescribing errors during the intervention phase (Table 4). After the enhanced A&F intervention ended, the month-to-month trend in prescribing errors at admission increased by 6% (p-value=0.028). However, relative to the control arm, the experiment arm had 57.6% (p-value=0.015) less prescribing errors at admission post-study, and the trend in the experiment arm relative to the control arm post-study had a 8.1% (p-value=0.005) month-to-month reduction in the admission prescribing errors (Table 4).

**Table 4:** Secondary analysis of the ReGENT study intervention effect on reducing all Gentamicin prescribing errors at admission (Hospitals = 22)

|  |  |  |  |  |  |  |
| --- | --- | --- | --- | --- | --- | --- |
|  | Pre-intervention phase compared to<br>intervention phase |  |  | Intervention phase compared to post-intervention phase |  |  |
|  | IRR | 95% CI | p-value | IRR | 95% CI | p-value |
| (intercept)<sup>1</sup> | 0.199 | 0.127 to 0.311 | <0.001 | 0.211 | 0.110 to 0.403 | <0.001 |
| time<sup>2</sup> | 0.951 | 0.922 to 0.981 | 0.002 | 0.980 | 0.950 to 1.011 | 0.195 |
| intervention<sup>3</sup> | 1.096 | 0.922 to 1.302 | 0.301 | 1.112 | 0.934 to 1.322 | 0.232 |
| intervention × I(time - 12)<sup>4</sup> | 1.090 | 1.036 to 1.146 | <0.001 | 1.060 | 1.006 to 1.116 | 0.028 |
| arm<sup>5</sup> (Ref: Control arm) | 0.769 | 0.470 to 1.325 | 0.371 | 0.424 | 0.213 to 0.847 | 0.015 |
| time × arm<sup>6</sup> | 1.032 | 0.998 to 1.067 | 0.064 | 1.042 | 1.009 to 1.077 | 0.011 |
| arm × intervention × I(time - 12)<sup>7</sup> | 0.930 | 0.878 to 0.985 | 0.014 | 0.919 | 0.866 to 0.975 | 0.005 |
| **Note:**<br><sup>1</sup>Baseline performance at the start of the pre-intervention period<br><sup>2</sup>Pre-intervention trend<br><sup>3</sup>Step change in performance due to the intervention<br><sup>4</sup>Change in the trend post-intervention<br><sup>5</sup>Baseline performance in the experiment arm relative to the control arm<br><sup>6</sup>Pre-intervention trend in the experiment arm relative to the control arm<br><sup>7</sup>Change in the trend post-intervention in the experiment arm relative to the control arm |  |  |  |  |  |  |

### Fidelity to intervention plan/ participants explanations for observed effects

Feedback about the draft findings of the ReGENT study was elicited from the paediatricians and nurses in charge of the newborn units of the hospitals that received the intervention. A key limiting factor was the small number of pharmacists and paediatricians available in the whole hospital vis-à-vis the numbers of neonatal patients and the number of junior clinicians (including interns) that constrained the amount of time they had to provide mentorship on antimicrobial stewardship to junior clinicians. The mentorship challenge was further exacerbated by the high staff turnover coupled with frequent rotation of interns (who provide the bulk of the patient care) every 8 – 12 weeks that was unsynchronised across the medical and nursing carders making it difficult to sustain any departmental progress in practice. There was clear indication that in some hospitals, the practices being reinforced were in conflict with what was in the clinical guidelines e.g. use of admission weight instead of birth weight to calculate the dosage to prescribe. Additionally, there were only six unique downloads and use of the Android-based smartphone application providing interactive feedback during the study duration.

## Discussion

### Summary of findings

Applying a theory-informed A&F intervention, the ReGENT study found no statistically significant effect of the team-based pharmacist-led A&F intervention on reducing gentamicin medication errors in neonatal care. Prescribing errors during intervention and post-intervention periods were increasing across all hospitals in both arms of the study. However, relative to control hospitals sites receiving routine feedback but without pharmacist involvement or pharmacist-based CME, the ReGENT study sites had a 7.3% less Gentamicin prescription error rates at admission post-introduction of the pharmacist-led A&F intervention, and 8.1% less Gentamicin prescription error rates at admission in the post-intervention phase.

### Comparison to other studies

The Gentamicin prescribing error-rates in neonatal care settings of 11%-26.4% at the beginning this study are consistent with previously published studies [9]. Studies using pharmacist-led A&F interventions have demonstrated at 9.6% drop in hazardous prescribing after 12 months, whereas from the findings in our secondary analysis, error rates during intervention and post-intervention periods were increasing across all places but more so in the control hospital sites, where they were significantly higher [15]. Key similarities between our study and other studies remained the use of providing actionable summaries linked to patient-level data that could be used to review patient care retrospectively. There was a fundamental difference between the previous studies where pharmacists reviewed individual at-risk patients, initiating remedial actions or advising doctors to do so. In our study, pharmacists were too few (typically one per hospital) and could not provide individual patient prescription reviews; They were constrained to co-ordinating trainings through CMEs for the clinical teams doing the actual prescribing and reviewing together with the neonatal teams organisational challenges impeding improvement of medication prescribing accuracy (e.g. medication stock-outs, lack of training etc.). In typical Kenyan clinical settings, there is little interaction between clinical teams and pharmacists to guide medication prescribing with medications reconstituted and administered by nurses on the ward; pharmacists typically only get involved for inpatient care when potentially toxic medications (e.g., chemotherapy) are administered but this is a rare event confined to higher level hospitals. Before the onset of this study, pharmacists had not previously been involved in CIN feedback activities except in some hospitals linked to the “*Supportive care and antibiotics for severe pneumonia among hospitalized children (SEARCH)*” trial [45] where their role was to support correct use of study drugs used in the paediatric wards. In this study, while pharmacists did not start routinely visiting the NBUs, they became more involved in the CMEs going through the ReGENT feedback reports.

### Interpretation of study findings

Our study findings indicates that the theory-driven pharmacist-led enhanced A&F intervention did not significantly change the trend in Gentamicin prescribing practices in the participating hospitals, with the overall level of prescribing errors getting worse across all hospitals. Compared to the control sites, in the ReGENT sites the trend in Gentamicin prescribing errors at admission during- and post-intervention was relatively better, which might be due to other events outside the study that might be affecting the provision of inpatient neonatal care (e.g. perhaps new clinicians trained during COVID had less idea what to do on NBUs); The control sites, defined by pharmacists declining to take part in the study, may suggest a lower level of engagement with, and the value placed on, NBU prescribing and care by hospital pharmacists.

The need for frequent and varied means of improving the awareness of current guidelines and care-giving norms around prescribing practices was also highlighted. Clinicians argued that the adherence to prescribing guidelines was further complicated where the newborns had other underlying complications – which is common hospitals in most SSA contexts-but the clinical guidelines offered no clear direction on what should happen in that scenario. However, the presence of the WhatsApp group allowed the pharmacists to seek peer support in what advice to offer hospital teams handling such cases. The use of visual aides at patient level (e.g. stickers on patient folders) at the point of care to flag where dosing errors would be critical, involving nurses in advising junior medical doctors and physician assistants (i.e. clinical officers) on the correct practices before prescription administration, and the use of multi-disciplinary induction and ward-round approaches to mentorship were emphasised as means to improve the prescribing practice. While the clinicians argued that they would rapidly respond to and act-on feedback provided through digital platforms, from the almost negligible download and use of the Android-based smartphone application providing interactive feedback, we saw a huge disconnect between the perception and the actual practice that A&F digital platforms would improve access and engagement with feedback, even where the such digital platforms were co-designed with clinicians.

### Study strengths, limitations and generalisability of findings

A key strength of this study was the natural control group that was used in the secondary analysis which was able to mitigate against the potential of history bias (a primary threat to the validity of in single-arm ITS studies) where concurrent interventions or events occurring around the time of the intervention in the facility [44]. Also, we reported patterns and trends in absolute error rates by patient sub-groups which are important results that are rarely reported from SSA studies. A key limitations of this study was the inability of the pharmacists to engage in individual patient reviews (the ideal situation), due to being overstretched (in terms of ongoing typical facility-based tasks) coupled with the high neonatal inpatient admissions. Additionally, the high staff turnover happening every 2-3 months made it difficult to sustain any improvements made through mentorship leading to a stagnation of the prescribing errors at admission. Methodologically, this study did not have sufficient statistical power to allow for ITS analyses at the birthweight and age sub-groups (i.e. the 3mg, 5mg or 7.5mg dosing levels), or the error types sub-groups (i.e. underdose, overdose). However, the findings of this study can be used to inform future pharmacist-led team-based interventions where resources are limited, the teams are large, and there is a high staff turnover.

## Conclusions

We found no statistically significant effect of the team-based pharmacist-led A&F intervention on reducing gentamicin medication errors in neonatal care. Prescribing errors during intervention and post-intervention periods were increasing across all hospitals in both arms of the study during and post-intervention periods. However, relative to control hospitals sites receiving routine feedback but without pharmacist involvement or pharmacist-led CMEs, the primary study sites had a positive trend in reducing Gentamicin prescription error rates at admission during and post-introduction of the pharmacist-led A&F intervention which might be due to other events outside the study that might be affecting the provision of inpatient neonatal care.

## Abbreviations

A&F: Audit and Feedback
CIN: Clinical Information Network
CME: Continuous Medical Education
CP-FIT: Clinical Performance Feedback Intervention Theory
EHRs: Electronic Health Records
HICs: High Income Countries
HCWs: Health Care Workers
ITS: Interrupted Time Series
KPA: Kenya Paediatric Association
LMICs: Low and middle-income countries
MoH: Ministry of Health
NBU: Newborn Unit
QI: Quality Improvement
SERU: KEMRI’s Scientific and Ethics Review Unit
SSA: Sub-Saharan Africa
WHO: World Health Organization.

## Declarations

### Ethics approval and consent to participate

Ethical approval was provided by the KEMRI Scientific and Ethical Review Committee (SERU 4378 and SERU 3459). The Scientific and Ethics Review Unit of the Kenya Medical Research Institute (KEMRI) approved the collection of the de-identified anonymised data for this study waiving the need for individual consent for access to de-identified patient data, with the authors having no access to the information that could identify the patients.

### Consent for publication

This study is published with the permission of the Director of Kenya Medical Research Institute (KEMRI).

### Availability of Data and Materials

The datasets generated and/or analysed during the current study are not publicly available due to the primary data being owned by the hospitals and their counties with the Ministry of Health; The research staff do have permission to share the data without further written approval from both the KEMRI-Wellcome Trust Data Governance Committee and the Facility, County or Ministry of Health as appropriate to the data request.

Requests for access to primary data from qualitative research by people other than the investigators will be submitted to the KEMRI-Wellcome Trust Research Programme data governance committee as a first step through, who will advise on the need for additional ethical review by the KEMRI Research Ethics Committee.

## Funding

This work was primarily supported by a Wellcome Trust Senior Fellowship (#207522/Z/17/Z) awarded to ME and a Wellcome Trust Early Career Research Fellowship (#227562/Z/23/Z) awarded to TT. Additional support was provided by a Wellcome Trust core grant awarded to the KEMRI-Wellcome Trust Research Programme (#092654). The funders had no role in the preparation of this report or the decision to submit for publication.

### Author contributions

Authorship eligibility guidelines for the final reports adhere to the Contributor Roles Taxonomy (CRediT) statement guidelines.

Conceptualization: TT, ME; Supervision: TT, JA; Methodology: TT, MM, DA, MO, JA; Formal Analysis: TT, MM, JA, ME; Investigation: TT, JA, ME; Resources: TT, ME, DA, MM; Data Curation: TT, GM; Writing – original draft: TT, ME; Writing – review & editing: TT, MO, JA, DA, MM, GM, ME; Funding acquisition: TT, ME;

## Data Availability

The datasets generated and/or analysed during the current study are not publicly available due to the primary data being owned by the hospitals and their counties with the Ministry of Health The research staff do have permission to share the data without further written approval from both the KEMRI-Wellcome Trust Data Governance Committee and the Facility, County or Ministry of Health as appropriate to the data request. Requests for access to primary data from qualitative research by people other than the investigators will be submitted to the KEMRI-Wellcome Trust Research Programme data governance committee as a first step through, who will advise on the need for additional ethical review by the KEMRI Research Ethics Committee

## Acknowledgements

The **Clinical Information Network (CIN) Group**: The CIN group hospital teams who are tagged to collaborate in the network’s development, data collection, data management, implementation of audit and feedback interventions and who will participate in this study include the following focal persons:

a. **Paediatricians:** Juma Vitalis, Nyumbile Bonface, Roselyne Malangachi, Christine Manyasi, Catherine Mutinda, David Kibiwott Kimutai, Rukia Aden, Caren Emadau, Elizabeth Atieno Jowi, Cecilia Muithya, Charles Nzioki, Supa Tunje, Dr. Penina Musyoka, Wagura Mwangi, Agnes Mithamo, Magdalene Kuria, Esther Njiru, Mwangi Ngina, Penina Mwangi, Rachel Inginia, Melab Musabi, Emma Namulala, Grace Ochieng, Lydia Thuranira, Felicitas Makokha, Josephine Ojigo, Beth Maina, Catherine Mutinda, Mary Waiyego, Bernadette Lusweti, Angeline Ithondeka, Julie Barasa, Meshack Liru, Elizabeth Kibaru, Alice Nkirote Nyaribari, Joyce Akuka, Joyce Wangari;
b. **Pharmacists:** Babra Murila, Beatrice Kamau, Caroline Naliaka, Cynthia Nduta, Evans Gitu, Evans Makumba, Esther Nduta, Lydia Momanyi, Matini Duncan, Sally Mugo, Sarah Kibira, Stephene Gichana, Rogers Omolo, Roy Mwendwa, Wycliffe Dunde
c. **Nurses:** Amilia Ngoda, Aggrey Nzavaye Emenwa, Patricia Nafula Wesakania, George Lipesa, Jane Mbungu, Marystella Mutenyo, Joyce Mbogho, Joan Baswetty, Ann Jambi, Josephine Aritho, Beatrice Njambi, Felisters Mucheke, Zainab Kioni, Jeniffer, Lucy Kinyua, Margaret Kethi, Alice Oguda, Salome Nashimiyu Situma, Nancy Gachaja, Loise N. Mwangi, Ruth Mwai, irginia Wangari Muruga, Nancy Mburu, Celestine Muteshi, Abigael Bwire, Salome Okisa Muyale, Naomi Situma, Faith Mueni, Hellen Mwaura, Rosemary Mututa, Caroline Lavu, Joyce Oketch, Jane Hore Olum, Orina Nyakina, Faith Njeru, Rebecca Chelimo, Margaret Wanjiku Mwaura, Ann Wambugu, Epharus Njeri Mburu, Linda Awino Tindi, Jane Akumu, Ruth Otieno, Slessor Osok;
d. **Health Record Information Officers (HRIOs):** Seline Kulubi, Susan Wanjala, Pauline Njeru, Rebbecca Mukami Mbogo, John Ollongo, Samuel Soita, Judith Mirenja, Mary Nguri, Margaret Waweru, Mary Akoth Oruko, Jeska Kuya, Caroline Muthuri, Esther Muthiani, Esther Mwangi, Joseph Nganga, Benjamin Tanui, Alfred Wanjau, Judith Onsongo, Peter Muigai, Arnest Namayi, Elizabeth Kosiom, Dorcas Cherop, Faith Marete, Johanness Simiyu, Collince Danga, Arthur Otieno Oyugi, Fredrick Keya Okoth.

The **Clinical Information Network (CIN) Group**’s monitored email address is and the list can change when new paediatrician(s), nurse(s) or HRIO leave or come into the hospital.

## Open access

This is an open access article distributed in accordance with the Creative Commons Attribution 4.0 Unported (CC BY 4.0) license, which permits others to copy, redistribute, remix, transform and build upon this work for any purpose, provided the original work is properly cited, a link to the licence is given, and indication of whether changes were made.

## Appendix

**Supplementary Table 1:** Pharmacist-led virtual Continuous Medical Education sessions (CMEs) convened during ReGENT study.

| No. | Topic | Date |
| --- | --- | --- |
| 1. | Introduction to gentamicin (mode of action, pharmacokinetics, adverse events, case study) | 26<sup>th</sup> July 2022 |
| 2. | Medication Safety/Medication Errors | 30<sup>th</sup> September 2022 |
| 3. | Appropriate antimicrobial use | 29<sup>th</sup> November 2022 |
| 4. | Medication Therapy Management | 27<sup>th</sup> March 2023 |

**Supplementary Table 2:** A&F intervention components as informed by CP-FIT theory.

| # | Intervention component | CP-FIT hypotheses*: Feedback interventions are more effective when... | Control Arm | Experiment Arm |
| --- | --- | --- | --- | --- |
| 1 | The pharmacists have proposed roles as QI champions/facilitators. They will be supported to conduct a preliminary session for orientating clinical interns into the study when they start their three-month rotation in paediatric and new-born wards. They will also encourage the nursing staff to identify prescription dosing errors and politely feed this back to the medical staff together with the paediatricians. They will help disseminate monthly reminders on dosing instructions during their physical interactions with NBU ward staff. | a) *Champions*: Supportive individuals in the organisation dedicated to making changes a success.<br>b) *Competing priorities*: Clinical teams have minimal additional responsibilities and/or competing priorities.<br>c) *Workflow fit*: Feedback and action fit alongside existing organisational and team ways of working.<br>These elements contribute to effectiveness by: promoting credibility of the feedback, limiting the resources needed to provide or act on feedback and employing social influence in support of a need for behaviour change. | ☐ | ☒ |
| 2 | The pharmacists will also conduct two-monthly routine Continuous Medical Education (CME) sessions and review the performance A&F summaries with the new-born unit team for 15 minutes in the monthly morbidity and mortality meetings for the whole team or any other suitable forum at the local hospital | a) *Knowledge and skills in clinical topic and quality improvement*: Feedback targets health professionals with greater capability in the clinical topic under focus.<br>b) *Source knowledge and skill*: Delivered by a person or organisation perceived to have an appropriate level of knowledge or skill.<br>c) *Delivery to a group*: Feedback delivered to groups of recipients.<br>These elements contribute to effectiveness by: relying on social influence to enhance feedback credibility and | ☐ | ☒ |

| # | Intervention component | CP-FIT hypotheses*: Feedback interventions are more effective when... | Control Arm | Experiment Arm |
| --- | --- | --- | --- | --- |
|  |  | acceptance, building HCWs knowledge and skills to facilitate action, and when emphasising a common goal, leveraging teamwork to target HCWs' perception, intention, and behaviour |  |  |
| 3 | The pharmacists will also be members of a WhatsApp group whose purpose is to facilitate conversations about prescription practices between fellow pharmacists in hospitals in the same study arm. The membership of this WhatsApp group is limited to pharmacists only. The WhatsApp group will be used to disseminate monthly reminders on dosing instructions to be shared with the rest of hospital-specific clinical team. | a) *Peer discussion*: Feedback encourages recipients to discuss their performance with peers.<br>This element targets feedback perception and intention, by leveraging social influence to break down feedback's complexity, and identify possible practice improvements. | ☐ | ☒ |
| 4*** | The pharmacists will also be members of an additional 'within hospital' WhatsApp group whose purpose is to facilitate conversations about prescription practices with their hospital's healthcare workers posted to the NBUs | a) *Delivery to a group*: Feedback is delivered to groups of recipients.<br>b) *Peer discussion*: Feedback encourages recipients to discuss their feedback performance with peers.<br>c) *Problem solving and teamwork*: Feedback supports recipients to identify and develop solutions to reasons for suboptimal performance.<br>d) *Action planning*: Feedback provides solutions to suboptimal performance (or support recipients to do so). | ☐ | ☐ |

| # | Intervention component | CP-FIT hypotheses*: Feedback interventions are more effective when... | Control Arm | Experiment Arm |
| --- | --- | --- | --- | --- |
|  |  | These elements target feedback's actionability by evaluating if practice context is compatible with the expected target goals. They promote perception and intention by leveraging social influence to break down feedback's complexity when identify possible practice improvements. |  |  |
| 5*** | An interactive digital application platform that is mobile-friendly and auto-updated monthly used to deliver the enhanced A&F report summaries. The content of the interactive A&F feedback platform will be made up of three visualisations** | a) *Automation*: Data collection and analysis is (near) automatic.<br>b) *Active delivery*: They “push” feedback messages to recipients rather than requiring them to “pull”.<br>c) *Usability*: Feedback delivered employs user-friendly designs.<br>d) *Performance level, timeliness, trend, and benchmarking*: Feedback uses recent data to communicate when recipients' current performance has room for improvement, how recipients' current performance in relation to their past performance and compares recipients' current performance to that of other NBUs.<br>e) *Importance, controllability, and relevance*: Feedback focuses on meaningful goals perceived to be in HCWs control and relevant to their role.<br>These elements seek to improve perception of and interaction with feedback, provide a relative advantage based on whether | ☐ | ☐ |

| # | Intervention component | CP-FIT hypotheses*: Feedback interventions are more effective when... | Control Arm | Experiment Arm |
| --- | --- | --- | --- | --- |
|  |  | the cost of deploying feedback is considered inexpensive in terms of time, human or financial resources. They also seek to promote social influence on (and perception of) feedback while solidifying its actionability, credibility and acceptance, and supporting intention for behaviour change. |  |  |
| 6 | Enhanced A&F soft-copy (PDF) infographic report generated monthly outlining the proportion of patients who received erroneous gentamicin prescriptions, delivered to the NBU team. These additional A&F reports will be delivered to both the hospital pharmacists, the consultant paediatrician or neonatologist in-charge of the neonatal unit, senior nurses, and the medical staff working on rotation in the unit for the duration of the study. | a) *Usability*: Feedback delivered employs user-friendly designs.<br>b) *Performance level, timeliness, trend, and benchmarking*: Feedback uses recent data to communicate when recipients' current performance has room for improvement, how recipients' current performance in relation to their past performance and compares recipients' current performance to that of other NBUs.<br>c) *Importance, controllability, and relevance*: Feedback focuses on meaningful goals perceived to be in HCWs control and relevant to their role.<br>These elements seek to improve perception of and interaction with feedback, provide a relative advantage based on whether the cost of deploying feedback is considered inexpensive in terms of time, human or financial resources. They also seek to promote social influence on (and perception of) feedback while |  | ☒ |

| # | Intervention component | CP-FIT hypotheses*: Feedback interventions are more effective when... | Control Arm | Experiment Arm |
| --- | --- | --- | --- | --- |
|  |  | solidifying its actionability, credibility and acceptance, and supporting intention for behaviour change. |  |  |

**Note:**
\* Concepts are expounded upon in detail elsewhere: (Brown, B., Gude, W.T., Blakeman, T. et al. *Clinical Performance Feedback Intervention Theory (CP-FIT): a new theory for designing, implementing, and evaluating feedback in health care based on a systematic review and meta-synthesis of qualitative research. Implementation Sci* 14, 40 (2019). https://doi.org/10.1186/s13012-019-0883-5)
\*\* Discussed in-depth in the study's published protocol [17]
\*\*\*Represents a deviation from the protocol where A&F intervention component was challenging to operationalise

**Supplementary Table 3:** Analysis of A&F intervention effect following intention-to-treat principles (Hospitals = 22)

|  | IRR | 95% CI | p-value |
| --- | --- | --- | --- |
| (intercept)<sup>1</sup> | 0.145 | 0.105 to 0.201 | <0.001 |
| time<sup>2</sup> | 0.980 | 0.958 to 1.003 | 0.090 |
| intervention<sup>3</sup> | 1.091 | 0.916 to 1.299 | 0.329 |
| intervention × I(time - 12)<sup>4</sup> | 1.024 | 0.989 to 1.061 | 0.181 |
| arm<sup>5</sup> (Ref: Control arm) | 1.315 | 0.836 to 2.067 | 0.236 |
| time × arm<sup>6</sup> | 0.988 | 0.959 to 1.016 | 0.392 |
| arm × intervention × I(time - 12)<sup>7</sup> | 1.016 | 0.967 to 1.068 | 0.533 |

**Note:**
<sup>1</sup>Baseline performance at the start of the pre-intervention period
<sup>2</sup>Pre-intervention trend
<sup>3</sup>Step change in performance due to the intervention
<sup>4</sup>Change in the trend post-intervention
<sup>5</sup>Baseline performance in the experiment arm relative to the control arm
<sup>6</sup>Pre-intervention trend in the experiment arm relative to the control arm
<sup>7</sup>Change in the trend post-intervention in the experiment arm relative to the control arm

---

**Supplementary Figure 1**: Breakdown of trends in Gentamicin prescribing errors at admission

**Supplementary Figure 2**: Hospital specific trends in Gentamicin prescribing errors at admission

**Supplementary Figure 3**: Hospital specific trends in Gentamicin prescribing overdose errors at admission

**Supplementary Figure 4**: Hospital specific trends in Gentamicin prescribing underdose errors at admission

